# Communicating sickle cell trait results after newborn screening: Approaches and implications to families

**DOI:** 10.1101/2022.10.30.22281739

**Authors:** Daima Bukini, Irene Msirikale, Emanuela Marco, Michael Msangawale, Lulu Chirande, Columba Mbekenga, Karim Manji, Julie Makani

## Abstract

**Introduction:** Tanzania is amongst the countries in Africa with one of the highest prevalence of individuals with Sickle Cell Trait (SCT). Identifying individuals with SCT is important as they may potentially have children with Sickle Cell Disease (SCD). Interventions such as Newborn Screening (NBS) for SCD can identify individuals carrying the gene very early on to explore strategies for primary prevention.

**Aim:** This study aims to document experiences and perspectives of families who have received SCT results for their children through the NBS Program. We were interested to learn their perspectives on the communication approaches used and implications of the results to families. Our overall goal is to evaluate what approaches works best to support comprehension, understanding of genetic testing, concepts of inheritability and general understanding of SCD. We further aim to explore key issues considered by families as most important to inform not only methods, but also most locally relevant content to guide genetic counselling sessions.

**Methods:** In total 29 families provided with SCT results participated in six (6) Focus Group Discussions. Families were recruited through NBS program implemented between June to September 2021. Analysis of the data was done through thematic content analysis.

**Results:** Findings were categorized into two main categories; (**1) Key issues to consider when communicating sickle cell trait results to families**. The following themes were identified under this category; (1a) Language used to explain the results (1b) Methods used to provide the results (1c) Who was provided with the results (1d) Families comprehension of the results and (1e) What influences families’ understanding of the results **(2) What are the implications of the results to families**. The following themes were identified under this category; (2a) How results influenced future reproductive choices (2b) How will the information be kept within families (2c) Age a child will start to be informed about the results (2d) How results influence gender blames within families.

**Conclusion:** Understanding how to ensure genetic results have been properly communicated is core in developing a genetic counselling program. In places where the programs are not well established, there is a need to explore contexts specific approaches to inform ethically relevant communication models that incorporated families and patient perspectives. This study un-packed the different aspects to consider when developing proper communication models and further highlighted issues to explore with families after receiving the results, with the hope that this information will help to inform genetic counselling sessions in places with high SCD burden.

## Introduction

The prevalence of sickle cell trait (SCT) in Africa has been reported to be high (Burnham-Marusich et al., 2016; Délicat-Loembet et al., 2014; Ndeezi et al., 2016; Obed et al., 2017). Tanzania is amongst the countries with high prevalence of SCD as well as SCT globally. The number of children born with SCD is estimated to be between 11,000-16,000 births per year, whilst estimates for those with SCT is between 13% to 50% in some ethnic groups (Ambrose et al., 2017; Ambrose, Smart, et al., 2018; Chami et al., 2019; Saidi et al., 2016). SCD in the country accounts for at least 5% of under-five mortality, however, NBS for SCD if complemented with comprehensive care services has shown to reduce under five mortality by up to 90% (Kuznik et al., 2016; Makani et al., 2011, 2018; Ohene-Frempong et al., 2005; Tluway & Makani, 2017; Tshilolo et al., 2008). There have been considerable efforts in Tanzania and other countries in Africa where the burden of SCD is high to implement NBS for SCD programs (Bukini et al., 2021; Nkya et al., 2019). Through newborn screening for SCD, children with SCD and those with SCT can be identified early on and through proper strategy for primary prevention with the families, there is a chance of reducing the number of children born with SCD in the region. This signifies the importance of understanding the best approaches to engage with families to ensure that families at risk of or already affected by the disease are fully informed of the risks involved and available interventions. The programs will also provide opportunities to explore appropriate genetic counselling approaches that will be feasible in this context. As a step towards informing the contexts appropriate counselling approaches, there is a need to understand how genetic results for both SCD and SCT are communicated to families and how the approaches used influence the intended outcomes. Some studies done in Africa shared concerns on the challenges of communicating genetic results to families and therefore hinderance to comprehension (Bukini, Mbekenga, Nkya, Purvis, & Parker, 2020). This concern is shared even in places with established genetic counselling programs. With exception of few countries in Africa, genetic counselling is still not very well-established field which exacerbates the challenges of communicating genetic information. This study aims to document experiences and perspectives of families who have received SCT and SCD results for their children through the Newborn Screening Program in Dar es Salaam, Tanzania. We were interested to learn perspectives on the communication approaches used and implications of the results to families. Our overall goal is to evaluate what approaches works best to support comprehension, understanding of genetic testing, concepts of inheritability and general understanding of SCD. We further aim to explore key issues considered by families as most important to inform not only methods, but also most locally relevant content to guide genetic counselling sessions.

## Methods

### Sampling methods and study participants

This study was implemented as part of NBS programs for SCD in an urban setting in Dar es Salaam region. Families provided with SCT results were recruited from the NBS program between June to September 2021 implemented in Amana Regional Referral Hospital (ARRH) and Temeke Regional Referral Hospital (TRRH).. Until September 2021, a total of 2218 children were screened. Out of which, 33 children had SCD, and 260 children had SCT. For this paper, the focus is to learn on the perspectives employed to communicate SCT results to families and its implications thereafter. Recruitment started with the nurse asking permission to the families to be contacted to participate in this study. The nurse provides general information to the families about the project and those who were willing to be contacted were followed up with a telephone conversation and those agreed were consented to participate in this study. The recruitment methods has also been described elsewhere (Bukini, Mbekenga, Nkya, Malasa, McCurdy, et al., 2020)

### Data Collection Methods and Instruments

We have used Focus Group Discussions (FGDs) to learn experiences from the families on the best communication approaches used when providing results for SCT after newborn screening for SCD. We conducted six (6) FGDs with families who were provided with SCT results. Participants of the FGDs include; Mothers (20), Fathers (3), Grandmothers (2) and other relatives (4). The detailed description of the composition per FGD has been summarized above in Figure 1 above. All discussions were done in Swahili. Interview and FGDs guide were structured to understand five different aspects (1) What information were communicated to families (2) What methods were used to communicate the results? (3) How much was understood from that communication (4) What influences understanding? (5) What were the responses within families after the results?

**Figure 1.**
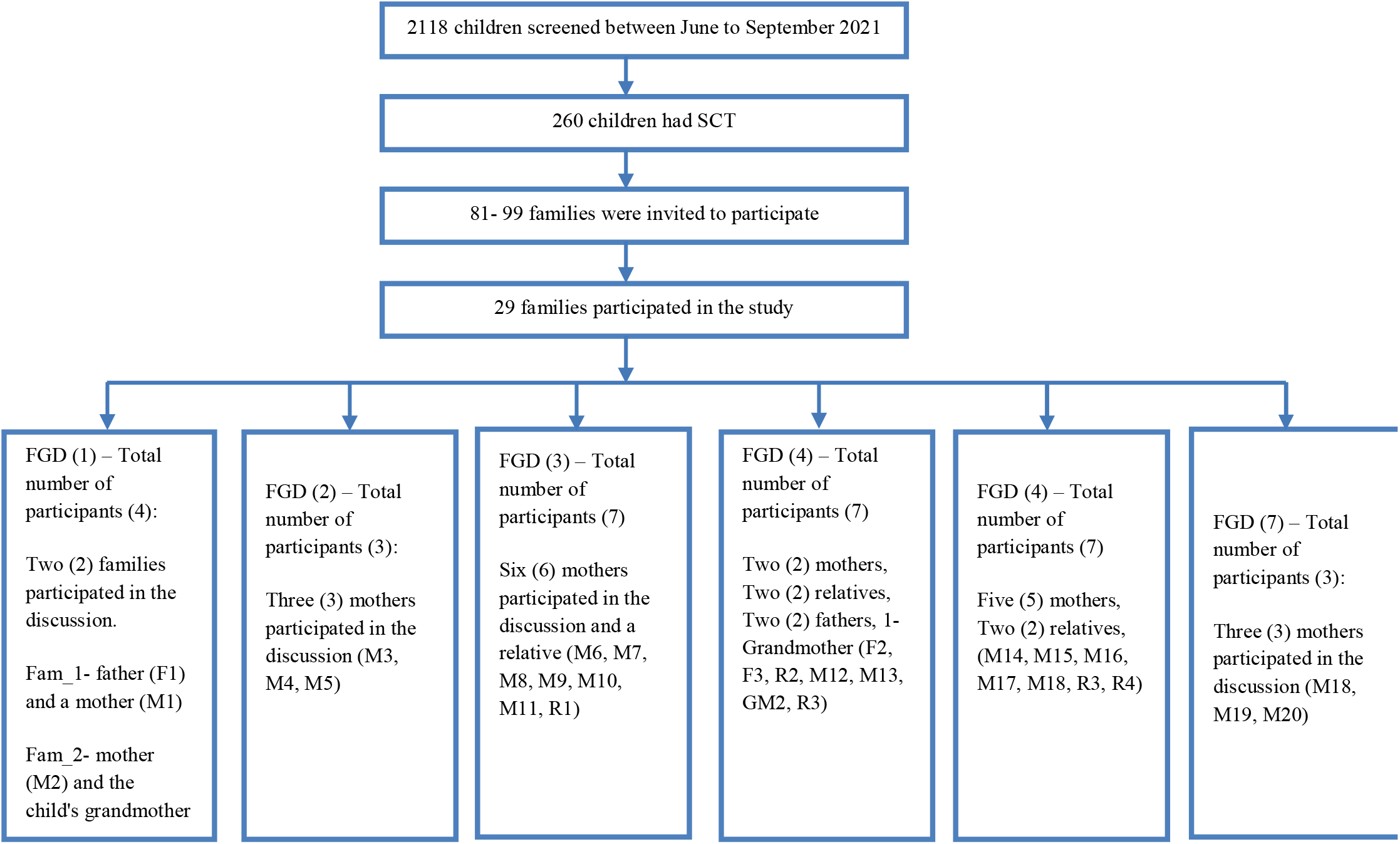
Recruitment flow chart for families provided with sickle cell trait results recruited through newborn screening program between June to September, 2021

**Figure 2:**
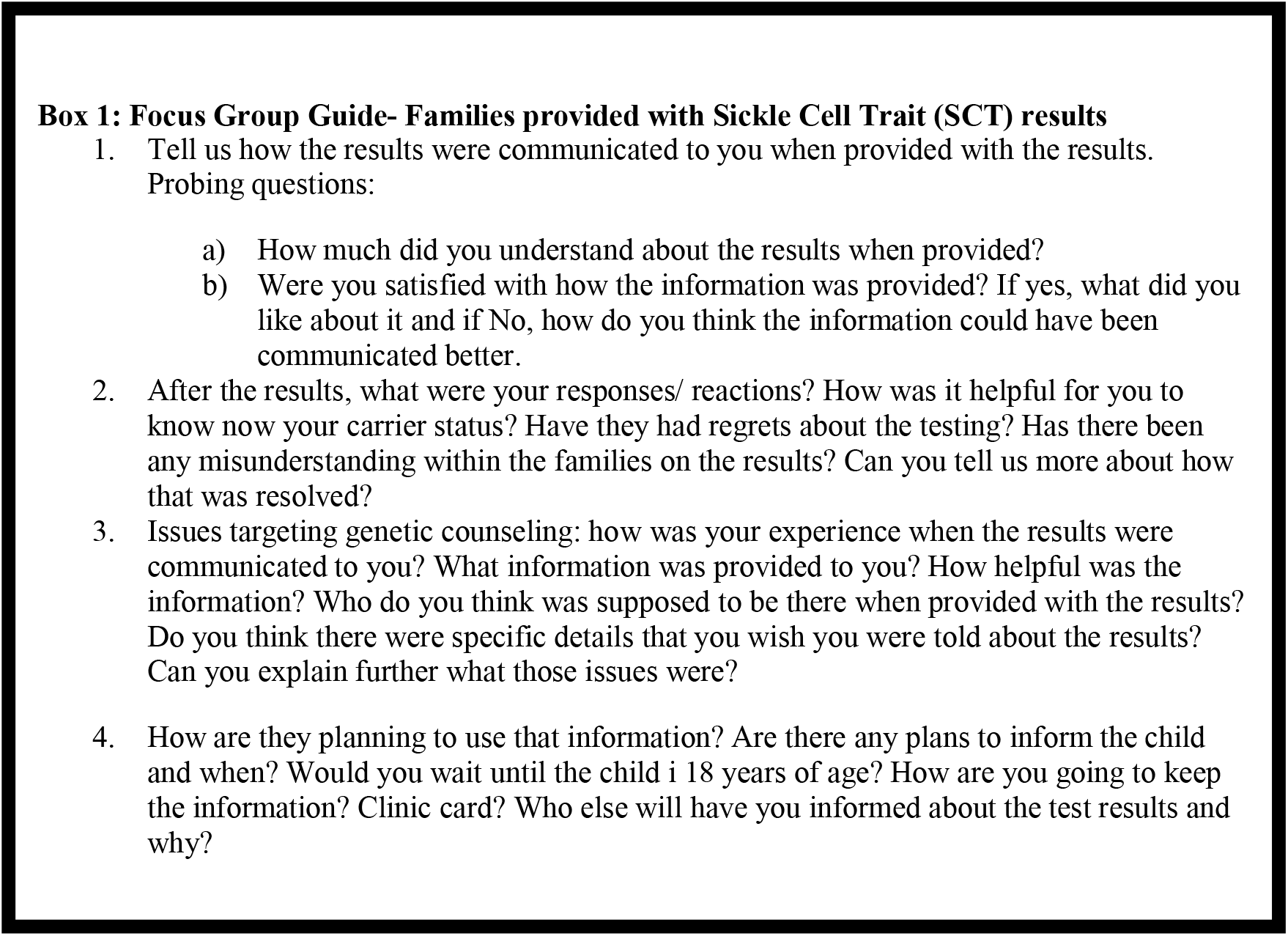
Sample of questions used for the Focus Group Discussions with families who participated in this study

### Data Analysis

All interviews and discussions were done in Swahili and audio recorded. DB transcribed the interviews and discussions and imported the transcripts into NVivo software and started coding of the interviews in Swahili. IM and EM double checked the coding to attain a certain level of consistency during coding to minimize subjectivity. All codes (not transcripts) were translated to English and used to develop a codebook. The codebook was tested by another group of researchers to test reliability and how robust it can be used to code the remaining transcripts. Meetings were held jointly to discuss the codebook and address any disagreement amongst the group. DB worked with IM and EM to identify connection of the different codes and identifying emerging themes that were later grouped into categories as summarized in the results section.

## Results

### 1.0 What were the key issues to consider when communicating results to families

There were several highlights during the discussions and interviews with the families that were considered as key when communicating results to families.

#### Language used to communicate the results

Specific examples provided here was how families interpreted differently the meaning of the word *‘genes*. There is more than one way in Swahili that translate to ‘genes’ and both of those words are not commonly used in swahili language-‘vinasaba’ and ‘chembechembe’. Families who were informed that your child has inherited SCT genes, with Swahili translation of ‘*chembechembe’* shown much better understanding than the translation to ‘*vinasaba’*. Summary of quotes below from FGD2_SCT families.

> *“For me ‘chembechembe’ was more comprehensible than ‘vinasaba’, because you know it is chembechembe it is not a complete thing (M4)… for me my child was told has ‘vinasaba’, and I did not understand, I had to ask (who provided the results), I was told don’t worry your child has small chembechembe for SCD, this is not going to cause any problem, but later when getting married and end up with a partner with ‘vinasaba’ like them, can get a child with SCD, but insisted that I should not be worried my child will not get sick, that’s when I understood the meaning” FGD2_Discussion with families provided with SCT results*

#### Methods used to communicate the results

Families provided with SCD results, a call was made to one of the parents, and they were requested to come to the clinic for the results. This provides an opportunity to have a one-to-one session with the clinicians. Families provided with AS results, communication was done through phone calls and for those who needed more information they were given an opportunity to come to the clinic. When asked which was the preferred methods of communication both groups indicated that a one-to-one session with the clinicians was important to provide them with opportunity to ask for more details.

> *“I received a call, honestly, I did not pick it up because the number was not familiar, I don’t know it, I was scared to pick the call. They call me three times. Later on, I decided to pick the call and listen, and that’s when (he/she) introduce themselves that they are doctor and ask if I recall my child being screened for SCD when born, I said yes, and (he/she) informed me that they found ‘chembechembe’ for SCD and ask if I can come to TRRH for more information. But when I was receiving the call, I was totally normal, I accept the situation and I wanted to seek help for my child and know more” FGD5_M16*.

#### Comprehension of the results

This was observed more in families provided with SCT results. Some of the parents did not seem to understand the distinction between having a child with SCD and SCT. Although explained during the phone call that their children do not have the disease, parents were still worried about children getting the disease later in their life.

> *“Ah…it has been almost a month since they call me, the first day when the results were out, they call me, they told me, your child has ‘vinasaba’ for SCD, and they told me when (the child) become adult and get a partner, they must go and get tested before living together. Because (they told me) there is a possibility of having a child with SCD. That is how they advise me, but also, they told me, when growing up, you must be careful (with the child) if start getting any complications like anaemia, and other health issues, you must send (the child) to a health facility. This is how they told me (M9)*.*…… For me I was very concerned when getting the results, (the doctor) told me it is just ‘vinasaba’ not the disease. But I was still worried, does it mean it is just a normal disease and not the real disease, the SCD, I was informed by the Doctor, between me and the father, one of us has vinasaba, And that’s how it went, I told them I will go to the hospital and listen what is going to happen to the child (M2)” FGD3_Discussion with SCT families*

#### Who was provided with the results

Procedurally, a call is made to either of the parents or a relative whose phone number is listed during the screening to provide the results. Some of the mothers shared their experiences of how shocked and confused they were when provided with the results. The suggestion provided was the results to be provided first to a family member (as preferred by the mother) who is going to share the news to the mother at a more convenient time. A good experience was when the results were shared to grandmother, who later communicated the massage to her daughter, although the mother was shocked and panicked but the grandmother was able to manage her situation very well.

> *“Even me when I received the results and heard the word ‘sickle cell’ I had to calm down and listen very carefully, fortunately they call me and not the mother and at that time I was not at home, I was farming. In the evening when I returned home, I told (the mother) I have received a call from the doctors, and they told me this and that (okay). The good thing when the child was born, she (the mother) informed me about the sickle cell screening, and at that time she (the mother) was so confused because her child was sick and she (mother) is very young (indicating that couldn’t cope well), she (the mother) put my number instead of her number” FGD 1_GM1*

#### What influenced understanding of the results

Families understanding was more influenced by either prior history within family of SCD or through knowing someone with SCD outside the family, either a friend or a neighbour. Families who already had a child with SCD were more informed about the disease and management than those with no prior history of the disease. In one of the discussions with AS families, one of the father recalls donating blood for his neighbour’s child who had SCD. He did recall how painful the experience was to both the child and the parents, and this experience helped him to seek for more information about the disease. In some cases, hearing or seeing someone with SCD within the community was a catalyst to ask for more information about the disease, compared to those who have heard about the disease through the screening program.

> *“For me, the way I understand sickle cell, is through seeing some people or may be in hospitals and another example is through TVs, to be honest, I don’t like seeing them. You will see a child stunted and very sick, even an adult looks like a child because of this problem, it is very big. It is painful. But, after some time (the doctors) tried to calm me down and explaining to me. (The doctor) told me it is not because it is a disease then you are going to lose your child (No), the more (the child) grow, we will be following up with the clinic and (the child) will live). But later, when they become adult (as explained by another participant) and wants to get married, before that they must get tested of the ‘vinasaba’ between the mother and the father, meaning the bride and the groom, and when they get the results, they will be able to plan accordingly” FGD3_M6*

#### 2.0 What were the potential next steps after the results and implications of the results to families

##### How are we going to keep the information

This was more important for families provided with SCT results since the results have an effect to the child’s offspring and not the child itself. The parents had lengthy discussions on what are the appropriate ways of storing the results within families until when the child will be able to comprehend the results. Although this was not agreed by everyone who has participated in the discussions but having hospital cards showing the child has AS was one way of keeping the information to show the child in the future or for other members to know. Other suggestions were for the information to be shared within the families to ensure that the child will be informed about the results in the future.

> *“I advise for families to be given special cards related to sickle cell, and they have to be special not with the clinic cards, because they have a lot of stuff. And you know ‘Mswahili’ when you see a lot of word, you don’t read at all, so I think for the sickle cell card to only be sickle cell card. For me that is my advice (M3)……*.*people within families are supposed to be informed, even when the child is growing up, you may not be there when the child is growing up, you can write on a piece of paper, we are human beings. Even those you have informed might not be there and no one will know if the child has a problem. So, I think you should write on a piece of paper and keep in a special place, when you are looking around the house, anyone can see (the paper) (M4)” FGD2*

##### At what age should we start involving the child

There was no age that the parents agreed to be more appropriate for the child to be informed because of how differently children behave at different age. They both agreed that the appropriate should be decided by the family on case-by-case basis depending on the child’s maturity. However, the most appropriate age was agreed to be between 13-14 years of age.

> *“I think starting from 13 to 14 years of age, at this age the child is mature enough, if you do quick calculations, the child might be in grade 7 au form 1 (secondary school), at this age, it is easy for them to understand (m0)…at this age they are very matured, they know what is good and what is bad, but for someone who is in grade 1, their level understanding is very low, but grade 7, only if the child is stupid but you can sit with the child and explain very well (about the results)” FGD1_Discussion with families provided with SCT results*

##### How the results influenced future reproductive choices

All the parents involved in this study did not know about their genotype except for those who had prior children with SCD. Families suggested for the SCD screening tests to be offered to families with AS results to help identify if both parents are carrier for SCD and therefore help the parents to make future reproductive decisions. However, for most of the parents in both groups, it is not clear how the results will influence the decision to have children in the future because for most of them that decision was largely informed by religious values and beliefs.

> *“You know the doctor is also a human being, if you tell me, you have this or that disease, those are only your words, not God’s words. First of all, how many people have the trait? How many of them are not even tested? What about you? Have you ever tested? All of you are not tested (pointing to other participants), but we are here, married and have children, it is only by Grace, for me there is a big lesson there, that’s why I am saying whose words are we following? A doctor? Who is also a human being?” FGD6_M20*

##### Gender blames within families

We did observe during the discussions that there were mothers who chose not to inform the fathers about the screening results, this was observed in both SCD and SCT groups. The main reason was they were worried of the children being stigmatized or the fathers abandoning the children because they either have SCD or SCT. Although, there was a case where a father was informed about the results (SCT) and chose not to inform the mother because he was concerned of how she is going to respond to the news.

> **“***The day when I received results, I call my husband, because he was not in town, he was very shocked, and he told me, he has never heard of such a thing. I told him those are the screening results, he said, mhh*.., *but you know he challenged me, he told me this is coming from your side” FGD6_M19*
>
> Table 1 summary of the results section highlighting the themes identified and examples of codes used in each of the themes 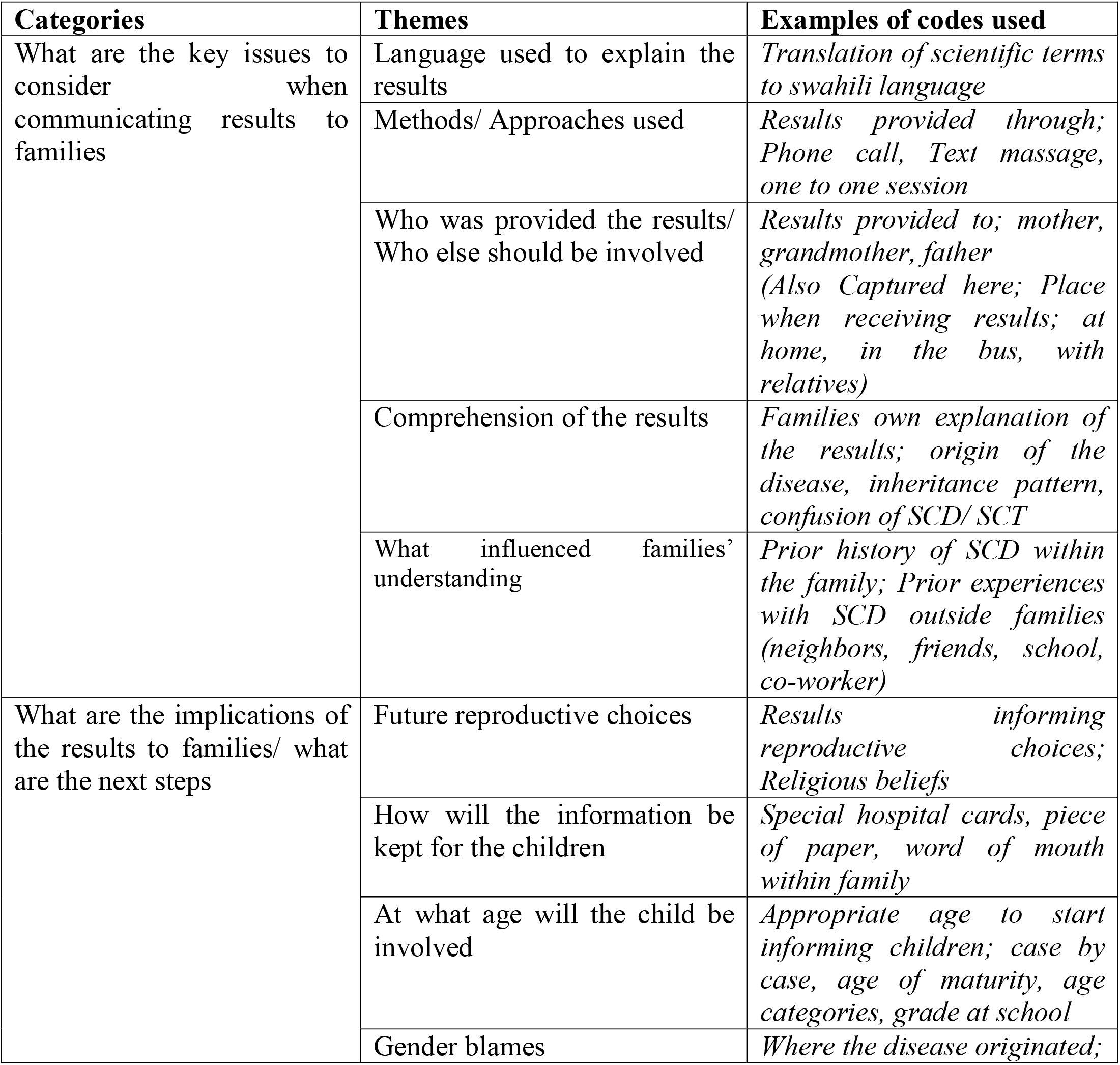

## Discussion

This paper is reporting on an ethically important question, how do we know we have <u>properly</u> communicated genetic results to families. We have brought together strong accounts and recommendations from the interviews and discussions done with families on what are the key issues to consider when communicating SCT results. Families understanding of the results is of priority especially in places where the burden of the SCD is high (Ambrose, Makani, et al., 2018; Fanning, 2016; Ndeezi et al., 2016). Results from this study has shown that families understanding of the results provided is still challenging. This observation suggests that despite the high birth prevalence of SCD in the area, still knowledge on the disease is very low (Abioye-Kuteyi et al., 2009; Ezenwosu et al., 2015; Obed et al., 2017; Tutuba et al., 2022). One of the recommendations to address this is to increase public awareness through cultural respectful advocacy campaigns within the communities (Anie et al., 2016; Brown, 2012). This may help to increase baseline knowledge of SCD amongst community members before they show up for the screening. Another key observation from the discussions, especially with families provided with SCT results was how they have misunderstood the implications of having a child who is a carrier for SCD. For most of them they thought the child has the disease and more interested to understand whether the child will start showing symptoms of the disease later in her life. This reflection was exacerbated by the limited knowledge on SCD within our communities, and the inadequate education provided before screening (Aboagye et al., 2019). Identifying catchment levels for provision of well-tailored educational programs to educate mothers on SCD and SCT is important. This will ensure mothers receive adequate education on the SCD and SCT and do understand implications of having a child with either of the two. Others suggested to start providing the health education programs during antenatal clinics where uptake/ attendance is very good and great possibility of finding both parents (Tutuba et al., 2022). Others have suggested counselling and education programs to be offered soon after screening (Ojodu, 2014). The language and the methods used to communicate were considered not fitting for the families. Similar studies done in Africa for genomic research reported similar findings (Kasule et al., 2022). The current practice is to provide results for SCD at the clinic and for the SCT, results are provided through telephone. Families recommended to have several levels of communication a) Telephone conversation with the family and b) face to face meeting with the clinician to allow them opportunity to ask questions and seek clarifications. Given the high prevalence of SCT in the area, having all families provided with SCT results at the clinics may not be feasible. One of the suggestions to address this can be to have them as a group to reduce time spent with individual families in the one to one sessions, somethings families were happy about (Bukini, Mbekenga, Nkya, Purvis, McCurdy, et al., 2020). Although the results were communicated in swahili but still there were terms which were new to families like *‘vinasaba’* a proper swahili translation for *‘genes*. Most of the families heard about this Swahili word for the first time when provided with the results, and for those who receive the results through telephone it was difficult for them to comprehend the meaning of the word. The most preferred word was *‘chembechembe’* a Swahili translation for ‘fine-particles’ to explain *‘genes*. ‘*Chembechembe’* is a common term used routinely and therefore families consider the term as a good metaphor to explain genes. Another example was SCD which is properly translated as *‘selimundu’* in swahili, but the most commonly used name is still ‘*sickle cell’* pronounced as *‘sikoseli’* in swahili. Although the two groups involved in this study were relatively distinct, there were common themes in both groups. For example, while gender blames is highly common amongst mothers with SCD children, mothers who have given SCT are also worried about the same issues. For some families the husbands or fathers of the children believed that the trait or disease has originated from the mothers family (Bukini, Mbekenga, Nkya, Malasa, McCurdy, et al., 2020). Future reproductive choices were also explored in detail amongst both groups. Based on the observation from this study it was not clear how having a child with SCD or SCT will influence future reproductive choices for the families. Perhaps, this was more influenced by the limited understanding of the concepts of heritability as shown by the families or religious believes more than having SCT/ SCD (Ibidunni et al., 2019; Pecker & Naik, 2018). While for those provided with SCD results, although parents were not tested, it is likely that both parents are carriers. However, for the SCT families, this was not established because the trait may either be coming from the mother or the father. One of the recommendations provided by the parents was for both parents whose children have been found to have SCT to be tested, to help inform their future reproductive choices (Burnham-Marusich et al., 2016). There was no age that was agreeable as the standard age to inform the child either of her carrier or disease status. Both families thought that the child should be involved based on their maturity on case-by-case basis because children of the same age may show different levels of maturity when involved in their health care (Borry et al., 2009). The discussion on how and where the results should be kept, ensuring that child will be made aware of their status as well as involvement of other family members.

## Conclusion

Understanding how to ensure genetic results have been properly communicated is core in developing a genetic counselling program. In places where the programs are not well established, there is a need to explore contexts specific approaches to inform ethically relevant communication models that incorporated families and patient perspectives. This study un-packed the different aspects to consider when developing proper communication models and further highlighted issues to explore with families after receiving the results, with the hope that this information will help to inform genetic counselling sessions in places with high SCD burden.

## Data Availability

All data produced in the present work are contained in the manuscript

## Ethical Consideration

The study was approved by the Muhimbili University of Health and Allied Sciences Research Ethics Committee (Ref. No. 2017-10-20/ AEC/ Vol. XII/85). Consent was sought prior to the conduct of the study

## Acknowledgements

We thank parents and caregivers who have participated in this research. The Sickle Cell Programme at Muhimbili University of Health and Allied Sciences. We also thanks staff and administration of Regional Referral Hospitals in Dar es Salaam.

## Authors contribution

DB designed the study, interacted with participants, performed interviews, assisted in transcriptions and translations, analysis and writing the first draft of the manuscript. KM, CM and JM supervised DB and reviewed the drafts of the manuscripts. IW assisted with data collection and reviewing the manuscript. All authors reviewed and approved the final manuscript.

## Data Availability

All data relevant to the study are included in the article

## REFERENCES

Abioye-Kuteyi, E. A., Oyegbade, O., Bello, I., & Osakwe, C. (2009). Sickle cell knowledge, premarital screening and marital decisions among local government workers in Ile-Ife, Nigeria. African Journal of Primary Health Care and Family Medicine. https://doi.org/10.4102/phcfm.v1i1.22

Aboagye, S., Torto, M., Asah-Opoku, K., Nuamah, M. A., Oppong, S. A., & Samba, A. (2019). Sickle cell education: A survey of antenatal healthcare givers. American Journal of Tropical Medicine and Hygiene, 101(3), 684–688. https://doi.org/10.4269/ajtmh.18-0408

Ambrose, E. E., Makani, J., Chami, N., Masoza, T., Kabyemera, R., Peck, R. N., Kamugisha, E., Manjurano, A., Kayange, N., & Smart, L. R. (2018). High birth prevalence of sickle cell disease in Northwestern Tanzania. Pediatric Blood and Cancer, 65(1). https://doi.org/10.1002/pbc.26735

Ambrose, E. E., Smart, L. R., Charles, M., Hernandez, A. G., Hokororo, A., Latham, T., Beyanga, M., Tebuka, E., Kamugisha, E., Howard, T. A., & Ware, R. E. (2018). Geospatial Mapping of Sickle Cell Disease in Northwest Tanzania: The Tanzania Sickle Surveillance Study (TS3). Blood. https://doi.org/10.1182/blood-2018-99-113939

Ambrose, E. E., Smart, L. R., Hokororo, A., Charles, M., Beyanga, M., Hernandez, A. G., Howard, T. A., & Ware, R. E. (2017). Prevalence and mapping of sickle cell disease in northwestern Tanzania. In Blood Advances. https://doi.org/10.1182/bloodadvances.2017GS101607

Anie, K. A., Treadwell, M. J., Grant, A. M., Dennis-Antwi, J. A., Asafo, M. K., Lamptey, M. E., Ojodu, J., Yusuf, C., Otaigbe, A., & Ohene-Frempong, K. (2016). Community engagement to inform the development of a sickle cell counselor training and certification program in Ghana. Journal of Community Genetics, 7(3), 195–202. https://doi.org/10.1007/s12687-016-0267-3

Borry, P., Evers-Kiebooms, G., Cornel, M. C., Clarke, A., & Dierickx, K. (2009). Genetic testing in asymptomatic minors: Background considerations towards ESHG Recommendations. European Journal of Human Genetics. https://doi.org/10.1038/ejhg.2009.25

Brown, S.-E. (2012). Cultural models of genetic screening & perceptions of sickle cell disease in high-risk Guadeloupean French communities [ProQuest Information & Learning]. In Dissertation Abstracts International Section A: Humanities and Social Sciences (vol. 72, Issues 9-A). http://ezproxy.ecu.edu.au/login?url=http://search.ebscohost.com/login.aspx?direct=true&db=psyh&AN=2012-99050-452&site=ehost-live&scope=site

Bukini, D., Mbekenga, C., Nkya, S., Malasa, L., McCurdy, S., Manji, K., Makani, J., & Parker, M. (2020). Influence of gender norms in relation to child’s quality of care: follow-up of families of children with SCD identified through NBS in Tanzania. Journal of Community Genetics. https://doi.org/10.1007/s12687-020-00482-4

Bukini, D., Mbekenga, C., Nkya, S., Purvis, L., McCurdy, S., Parker, M., & Makani, J. (2020). A qualitative study on aspects of consent for genomic research in communities with low literacy. BMC Medical Ethics, 21(1), 1–7. https://doi.org/10.1186/s12910-020-00488-0

Bukini, D., Mbekenga, C., Nkya, S., Purvis, L., & Parker, M. (2020). A qualitative study on aspects of consent for genomic research in communities with low literacy. 21(1), 1–7. https://doi.org/10.1186/s12910-020-00488-0

Bukini, D., Nkya, S., Mccurdy, S., Mbekenga, C., Manji, K., Parker, M., & Makani, J. (2021). Perspectives on Building Sustainable Newborn Screening Programs for Sickle Cell Diseasef□: Experience from Tanzania. 1–15.

Burnham-Marusich, A. R., Ezeanolue, C. O., Obiefune, M. C., Yang, W., Osuji, A., Ogidi, A. G., Hunt, A. T., Patel, D., & Ezeanolue, E. E. (2016). Prevalence of sickle cell trait and reliability of self-reported status among expectant parents in Nigeria: Implications for targeted newborn screening. Public Health Genomics. https://doi.org/10.1159/000448914

Chami, N., Hau, D. K., Masoza, T. S., Smart, L. R., Kayange, N. M., Hokororo, A., Ambrose, E. E., Moschovis, P. P., Wiens, M. O., & Peck, R. N. (2019). Very severe anemia and one year mortality outcome after hospitalization in Tanzanian children: A prospective cohort study. In PLoS ONE. https://doi.org/10.1371/journal.pone.0214563

Délicat-Loembet, L. M., Elguero, E., Arnathau, C., Durand, P., Ollomo, B., Ossari, S., Mezui-me-ndong, J., Mbang Mboro, T., Becquart, P., Nkoghe, D., Leroy, E., Sica, L., Gonzalez, J. P., Prugnolle, F., & Renaud, F. (2014). Prevalence of the Sickle Cell Trait in Gabon: A nationwide study. Infection, Genetics and Evolution, 25, 52–56. https://doi.org/10.1016/j.meegid.2014.04.003

Ezenwosu, O. U., Chukwu, B. F., Ikefuna, A. N., Hunt, A. T., Keane, J., Emodi, I. J., & Ezeanolue, E. E. (2015). Knowledge and awareness of personal sickle cell genotype among parents of children with sickle cell disease in southeast Nigeria. Journal of Community Genetics, 6(4), 369–374. https://doi.org/10.1007/s12687-015-0225-5

Fanning, J. B. (2016). Normative and Pragmatic Dimensions of Genetic Counselling: Negotiating Genetics and Ethics. Philosophy and Medicine, 124, 1637–1647. https://doi.org/10.1007/978-3-319-04414-9_126

Ibidunni, L., Nunez, K. P., Jonassaint, J. C., & De Castro, L. (2019). Regional Differences in the Beliefs and Practices Among Adults with Sickle Cell Disease Regarding Reproductive Health and Family Planning: A Sub-Analysis. Blood, 134(Supplement_1), 2114. https://doi.org/10.1182/blood-2019-129554

Kasule, M., Matshaba, M., Mwaka, E., Wonkam, A., & Vries, J. De. (2022). Considerations of Autonomy in Guiding Decisions around the Feedback of Individual Genetic Research Results from Genomics Researchf□: Expectations of and Preferences from Researchers in Botswana. 2022.

Kuznik, A., Habib, A. G., Munube, D., & Lamorde, M. (2016). Newborn screening and prophylactic interventions for sickle cell disease in 47 countries in sub-Saharan Africa: A cost-effectiveness analysis. BMC Health Services Research, 16(1). https://doi.org/10.1186/s12913-016-1572-6

Makani, J., Cox, S. E., Soka, D., Komba, A. N., Oruo, J., Mwamtemi, H., Magesa, P., Rwezaula, S., Meda, E., Mgaya, J., Lowe, B., Muturi, D., Roberts, D. J., Williams, T. N., Pallangyo, K., Kitundu, J., Fegan, G., Kirkham, F. J., Marsh, K., & Newton, C. R. (2011). Mortality in sickle cell anemia in africa: A prospective cohort study in Tanzania. PLoS ONE, 6(2). https://doi.org/10.1371/journal.pone.0014699

Makani, J., Tluway, F., Makubi, A., Soka, D., Nkya, S., Sangeda, R., Mgaya, J., Rwezaula, S., Kirkham, F. J., Kindole, C., Osati, E., Meda, E., Snow, R. W., Newton, C. R., Roberts, D., Aboud, M., Thein, S. L., Cox, S. E., Luzzatto, L., & Mmbando, B. P. (2018). A ten year review of the sickle cell program in Muhimbili National Hospital, Tanzania. BMC Hematology, 18(1), 1–13. https://doi.org/10.1186/s12878-018-0125-0

Ndeezi, G., Kiyaga, C., Hernandez, A. G., Munube, D., Howard, T. A., Ssewanyana, I., Nsungwa, J., Kiguli, S., Ndugwa, C. M., Ware, R. E., & Aceng, J. R. (2016). Burden of sickle cell trait and disease in the Uganda Sickle Surveillance Study (US3): a cross-sectional study. The Lancet Global Health, 4(3), e195–e200. https://doi.org/10.1016/S2214-109X(15)00288-0

Nkya, S., Mtei, L., Soka, D., Mdai, V., Mwakale, P. B., Mrosso, P., Mchoropa, I., Rwezaula, S., Azayo, M., Ulenga, N., Ngido, M., Cox, S. E., D’Mello, B. S., Masanja, H., Kabadi, G. S., Mbuya, F., Mmbando, B., Daniel, Y., Streetly, A., … Makani, J. (2019). Newborn screening for sickle cell disease: an innovative pilot program to improve child survival in Dar es Salaam, Tanzania. International Health. https://doi.org/10.1093/inthealth/ihz028

Obed, S. A., Asah-Opoku, K., Aboagye, S., Torto, M., Oppong, S. A., & Nuamah, M. A. (2017). Awareness of sickle cell trait status: A cross-sectional survey of antenatal women in Ghana. American Journal of Tropical Medicine and Hygiene, 96(3), 735–740. https://doi.org/10.4269/ajtmh.16-0396

Ohene-Frempong, K., Bonney, A., Tetteh, H., & Nkrumah, F. K. (2005). 270 Newborn Screening for Sickle Cell Disease in Ghana. Pediatric Research, 58(2), 401–401. https://doi.org/10.1203/00006450-200508000-00299

Ojodu, J. (2014). Incidence of Sickle Cell Trait — United States, 2010. Centers for Disease Control and Prevention MMWR. https://doi.org/10.1242/jeb.02455

Pecker, L. H., & Naik, R. P. (2018). The current state of sickle-cell trait: implications for reproductive and genetic counseling. Blood. https://doi.org/10.1182/blood-2018-06-848705

Saidi, H., Smart, L. R., Kamugisha, E., Ambrose, E. E., Soka, D., Peck, R. N., & Makani, J. (2016). Complications of sickle cell anaemia in children in Northwestern Tanzania. Hematology, 21(4), 248–256. https://doi.org/10.1080/10245332.2015.1101976

Tluway, F., & Makani, J. (2017). Sickle cell disease in AfricaflJ: an overview of the integrated approach to health, research, education and advocacy in Tanzania, 2004-2016. 177(6), 919–929. https://doi.org/10.1111/bjh.14594.Sickle

Tshilolo, L., Kafando, E., Sawadogo, M., Cotton, F., Vertongen, F., Ferster, A., & Gulbis, B. (2008). Neonatal screening and clinical care programmes for sickle cell disorders in sub-Saharan Africa: Lessons from pilot studies. Public Health, 122(9), 933–941. https://doi.org/10.1016/j.puhe.2007.12.005

Tutuba, H. J., Jonathan, A., Lloyd, W., Luoga, F., Marco, E., Ndunguru, J., Kidenya, B. R., Makani, J., Ruggajo, P., Minja, I. K., & Balandya, E. (2022). Prevalence of Hemoglobin-S and Baseline Level of Knowledge on Sickle Cell Disease Among Pregnant Women Attending Antenatal Clinics in Dar-Es-Salaam, Tanzania. Frontiers in Genetics, 13(April), 1–10. https://doi.org/10.3389/fgene.2022.805709

